# Efficient Deep Network Architecture for COVID-19 Detection Using Computed Tomography Images

**DOI:** 10.1101/2020.08.14.20170290

**Authors:** Chirag Goel, Abhimanyu Kumar, Satish Kumar Dubey, Vishal Srivastava

## Abstract

Globally the devastating consequence of COVID-19 or Severe Acute Respiratory Syndrome-Coronavirus (SARS-CoV-2) has posed danger on the life of living beings. Doctors and scientists throughout the world are working day and night to combat the proliferation or transmission of this deadly disease in terms of technology, finances, data repositories, protective equipment, and many other services. Rapid and efficient detection of COVID-19 reduces the rate of spreading this deadly disease and early treatment improve the recovery rate. In this paper, we proposed a new framework to exploit powerful features extracted from the autoencoder and Gray Level Co-occurence Matrix (GLCM), combined with random forest algorithm for the efficient and fast detection of COVID-19 using computed tomographic images. The model’s performance is evident from its 97.78% accuracy, 96.78% recall, and 98.77% specificity.

## 1. Introduction

The huge number of COVID-19 (coronavirus disease-19) cases has put the entire world in turmoil, leaving the researchers struggling to treat, track and possibly cure the disease. Laboratory tests are most accurate but they take time to report back, thus mass screening is not possible at a pace greater than the speed of corona spread. Contrary to the cases, there are only limited test kits available. Early identification is necessary because this helps the government to flatten the epidemic curve. Thus to avoid COVID-19 spread, it is important to incorporate automated fast and efficient detection method as an alternative diagnostic device before a significant clinical breakthrough is done for its cure which is proven and worldwide acceptable to the scientific community working in this domain.

For this purpose, deep learning based solutions have been proposed in literature [1]. Deep Learning is a subfield of machine learning that is influenced by the structure and function of the brain, known as artificial neural networks (ANN). It mainly focused on feature extraction, image classification and reconstruction. Employing the advances in Artificial Intelligence (AI) to the benefit of clinical decision making is becoming increasingly popular, especially in the battle against coronavirus [2]. AI involving medical imaging has been developed for image feature extraction, including shape and spatial perpetuity relation features [3]. A review on the use of AI in medical imaging can be found in [4]. Researchers have been peculiar to determine the possible limitations, and constraints of AI against COVID-19. Asnaoui et al. [5] conducted a comparative study of several recent deep learning models presented for the detection of coronavirus.

The coronavirus exhibits partially identical behavior to pneumonia, thus distinguishing and diagnosing becomes difficult to manage with the current rate of spreading rate. Hence, radiological imaging is a major diagnostic tool. Kassani et al. [6] used chest X-ray and computed tomography (CT) images to compare popular deep learning-based feature extraction frameworks by chosing MobileNet, DenseNet, Xception, ResNet, InceptionV3, InceptionResNetV2, VGGNet, NASNet to extract features which were then fed into several machine learning classifiers. Ardakani et al. [7] tested well-known convolutional neural networks like AlexNet, VGG-16, VGG-19, SqueezeNet, GoogleNet, MobileNet-V2, ResNet-18, ResNet-50, ResNet-101, and Xception; to evaluate their ability to distinguish COVID-19 from non-COVID-19 cases. Ilyas et al. [8] discussed architectures like ResNet, Inception, Googlenet, etc. and the challenges involved in the deployment of covid detection from chest X-ray images.

CT scans and X-ray images reveal the specific associated manifestations. It has been reported that bilateral and peripheral ground glass opacification (GGO) are predominant in CT findings of infected patients [9]. Chest CT is useful to assess the severity of lung infection [10]. Compared with reverse-transcription polymerase chain reaction (RT-PCR), chest CT images may be more trustworthy, useful, and possibly aid in rapid classification [11]. Pan et al. [12] analyzed the changes in chest CT findings of covid patients from initial diagnosis until recovery. Compared to the chest X-ray, the CT scans are widely preferred due to 3D view of the lung, which further provides 2D images of axial, coronal and sagittal-view for better diagnosis. Ozkaya et al. [13] stated that “Consensus occurred in the opinion that using CT techniques for early diagnosis of pandemic disease gives both fast and accurate results.” Wu et al. [14] studied the CT scans of infected patients to describe its relationship with clinical features, and confirmed that CT plays an important role in the diagnosis. Thus, deep learning methods will be able to extract graphical features nicely from CT scans to provide a clinical diagnosis ahead of the pathogenic test, thus saving critical time. Unfortunately, most of the earlier works published on this matter suffer due to unavailability of open-source dataset at that time, and were often stuck with very few images. A deep learning-based CT diagnosis system (DeepPneumonia) [15] was developed for identification purpose but employed small dataset. A deep learning framework COVIDX-Net was proposed in [16] for automatic diagnosis of COVID-19 from X-ray images which, however, was validated on a small dataset. Infact most of the work published in first three months of this year suffered due to training/testing on only tens or just a hundred images. Afshar et al. [17] presented an framework based on capsule networks which they suggested could even handle small datasets. However, methods like deep neural network are prone to lose spatial information between image instances and require large datasets. Even with this limitation of the early work, literature is enough to demonstrate the proof-of-principle that using learning-based methods on radiological graphics, one could develop a COVID-19 diagnostic system.

Most works are difficult to reproduce and adopt since the CT data used in such studies is not always publicly available. Besides such works employed small dataset in contrast to their real requirement of large CT image dataset. This was tackled by employing transfer learning (TL) technique [18]. Jaiswal et al. [19] proposed a TL approach on Pruned EfficientNet-based model for COVID-19 detection using chest radiographs and CT scans. TL serves as an effective mechanism in providing a solution by transferring knowledge from generic object recognition tasks to domain-specific tasks [20]. The performance of CNN architectures for medical image classification has been studied by adopting TL procedure [21, 22]. With TL, the detection of various abnormalities in small medical image datasets is achievable. Rajaraman et al. [23] employed knowledge transfer and iteratively pruned deep learning model ensembles for detection using chest X-ray images. A self-trans approach, which synergistically integrates contrastive self-supervised learning with TL to learn unbiased feature representations (for reducing the risk of overfitting) was proposed in [24]. However, in TL, the retention of knowledge extracted from one task remains the key to perform an alternative task. Currently, a school of researchers working in computer-aided medical research is investigating new architecture designs combined with clinical understanding for use in COVID-19 detection. Such architectures will have a long lasting impact as they will be useful in other applications as well, even after the pandemic is over.

For quantification of infectious regions, and fast manual delineation of training samples, a human-in-the-loop (HITL) strategy was proposed in [9] to assist radiologists. The average Dice similarity coefficient showed 91.6% agreement between automatic and manual segmentations. A deep learning based algorithm comprising of lung segmentation, 2D slice classification and fine grain localization was proposed in [25] to detect severity of manifestation from chest CT scans. A 2D segmentation model using the U-Net architecture gave output by segmenting the infectious region [26]. A random forest (RF) model was employed for the assessment of severity (non-severe or severe) of COVID-19 based on chest CT images [27]. Four pre-trained deep models (Inception-V3, ResNet-50, ResNet-101, DenseNet201) with multiple classifiers (linear discriminant, linear SVM, cubic SVM, KNN and Adaboost decision tree) were applied in [28] for severity identification from chest CT scans. A deep learning algorithm consisted of lesion detection, segmentation, and location was trained and validated on chest CT images for automatic detection of abnormalities [29].

For early phase detection, feature fusion and ranking method have been applied and then, the processed data was classified with a support vector machine [13]. Frequency domain algorithm, called FFT-Gabor scheme, was proposed in [30] to predict the patient’s state with an average accuracy of 95.37%. An architecture composed of an encoder and two decoders for reconstruction and segmentation was given in [31] which then employed multi-layer perceptron for classification based on chest CT images. Comparison of several CNN models for classification of CT samples into COVID-19, viral pneumonia, or no-infection has been conducted in [32]. For classification of covid infected patients using chest CT images, a CNN model was used where the obtained parameters were further tuned by multiobjective differential evolution [11].

Alom et al. [33] evaluated Inception Residual Recurrent CNN (with transfer learning approach) and NABLA-N network model on both X-ray and CT scan images. They demonstrated promising results for COVID-19 detection and infected region localization with respective models. Barstugan et al. [34] formed four different datasets by taking patch sizes from 150 CT images, where feature extraction was performed by Grey Level Co-occurrence Matrix (GLCM), Local Directional Pattern (LDP), Grey Level Run Length Matrix (GLRLM), Grey-Level Size Zone Matrix (GLSZM), and Discrete Wavelet Transform (DWT) algorithms, then SVM was used for classification. They reported best classification accuracy was obtained by GLSZM. Hasan et al. [35] proposed that each CT scan underwent a feature extraction involving deep learning and Q-deformed entropy algorithm, and then long short-term memory (LSTM) neural network classifier be used.

However, there are still certain limitations. Several researchers have themselves noted that network design and training can be improved. Also, the data employed in most studies did not perform cross-center validations. As the diagnostic algorithm is based on deep learning, so it works as a black-box which makes its explainability tougher. Some works partially tackled with one of the listed problems or a part of a problem. Related work of all limitations mentioned above will be addressed in our further studies. In the recent years, evolution of deep learning paradigm has made it possible to develop sophisticated automated feature extraction techniques, which enables efficient data compression into lower dimensions without significant information loss.

Advancement in the deep learning feature extraction methodologies makes it a potential tool especially in the field of medical imaging. An autoencoder is a neural network model, trained to reconstruct its input in an unsupervised way. Usually hidden network layers reduce the input size, and learn relevant features that allow better reconstruction. However, deep auto-encoders use several nonlinear layers to learn complex hierarchical functions from highly insightful results. To generate meaningful features particularly for image processing and natural language processing applications, approaches such as Sparse autoencoders [36], Denoising autoencoders [37], Contractive autoencoders [38] and Variational autoencoders [39] are. Nevertheless, these approaches only amount for data reconstruction.

In this paper, a new framework has been proposed which employs the extraction of two feature sets, the first feature set being extracted using an unsupervised learning approach, the convolutional autoencoders as generic feature extractor while the second set of features has been extracted, keeping in mind the importance of textural features for images, using Gray Level Co-occurrence matrix as hand-crafted feature to build a better performing classifier. Analysis have been performed on both sets of features to find out that the ensemble of both these feature sets can be useful for the classification of the SARS-CoV-2 when employed via a random forest classifier. Results showed that the proposed approach could be used to diagnose COVID-19 as an assistant framework. The details are discussed in the subsequent sections.

## 2. Methodology

Deep learning generally refers to application of CNN for feature extraction and object classification. The layers of CNN process information in a nonlinear fashion which transforms the data into a more abstract level. The neurons in a particular layer are selectively attached to some of the neurons in the next layer. Higher layers essentially enhance parts of the given data that are significant for segregation from unimportant attributes. Finally, the output is diminished to a single vector of probability scores. For COVID-19 infection detection, features of chest CT images are used for classification whether they belong to infected class or not.

### 2.1. Database

The dataset consists of 2482 CT scans, out of which 1252 images belong to patients tested positive with SARS-CoV-2 infection, and 1230 images for patients not infected with coronavirus (but infected with other pulmonary diseases). The dataset selected for the verification of proposed concept is already openly available.

### 2.2. Convolutional Autoencoder

Autoencoders, unsupervised learning generative models, are used to reconstruct the input image. They employ a symmetric model consisting of two blocks, viz. encoder and decoder. The encoder compresses input image into a lower dimension output that contains only the informative features of input, then the decoder reconstructs the image from the features extracted by the encoder. So once the training is completed the encoder becomes a powerful tool for the extraction of features from the input. These autoencoders can be created using different type of neural networks.

In this study, we employed a convolutional autoencoder architecture which is shown in Figure 1. In Table 1, the layers 1 to 9 constitute the encoder which operates on 224 × 224 × 3 pixels to convert it to a 512 feature vector, and the layers 10 to 18 constitute the decoder part which is useful for training but is not required for deployment.

**Figure 1:**
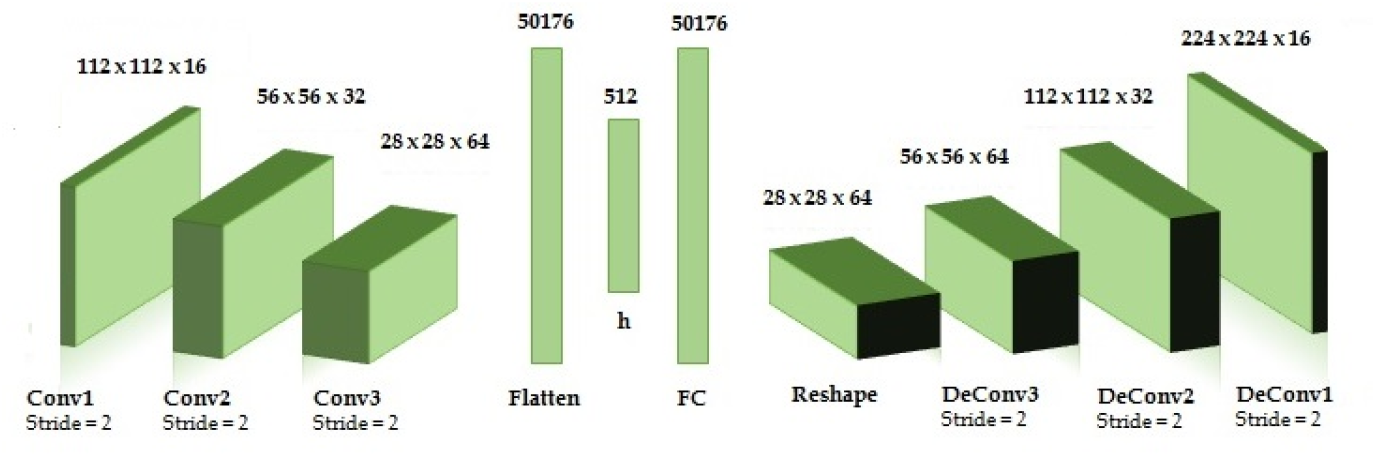
Flowchart of the proposed framework

**Table 1:** Description of autoencoder architecture layers

| Layer | Kernel Size | Width | Activation | Output Size |
| --- | --- | --- | --- | --- |
| Input | - | 3 | - | $224 \times 224 \times 3$ |
| Conv2D | $3 \times 3$ | 16 | ReLu | $224 \times 224 \times 16$ |
| max-pool | $2 \times 2$ | | | $112 \times 112 \times 16$ |
| Conv2D | $3 \times 3$ | 32 | ReLu | $112 \times 112 \times 32$ |
| max-pool | $2 \times 2$ | | | $56 \times 56 \times 32$ |
| Conv2D | $3 \times 3$ | 64 | ReLu | $56 \times 56 \times 64$ |
| max-pool | $2 \times 2$ | | | $28 \times 28 \times 64$ |
| Flatten |  |  |  | 50176 |
| Dense |  |  |  | 512 |
| Dense |  |  |  | 50176 |
| Reshape | | | | $28 \times 28 \times 64$ |
| Conv2D | $3 \times 3$ | 64 | ReLu | $28 \times 28 \times 64$ |
| up-sample | $2 \times 2$ | | | $56 \times 56 \times 64$ |
| Conv2D | $3 \times 3$ | 32 | ReLu | $56 \times 56 \times 32$ |
| up-sample | $2 \times 2$ | | | $112 \times 112 \times 32$ |
| Conv2d | $3 \times 3$ | 16 | ReLu | $112 \times 112 \times 16$ |
| up-sample | $2 \times 2$ | | | $224 \times 224 \times 16$ |
| Conv2D | $3 \times 3$ | 3 | Sigmoid | $224 \times 224 \times 3$ |

The objective here was to keep minimum possible features so the encoder can be tested with greater depth and multiple convolutional layers at each step. It was found that architecture of layers 1 to 9 gave the best results. Experimentation was conducted to compare 512 and 256 features, in which we found that one with 512 features performed better.

The network was trained using a batch size of 32 for a maximum of 200 epochs with Adam optimizer and mean squared error (MSE) as loss function. The model was trained using early stopping which ensured that if the validation loss remained non-decreasing consistently for 30 epochs then the training stopped. The objective remains to avoid overfitting of the model and storing the best model at model checkpoint by minimizing the validation loss.

### 2.3. Haralick Features

Haralick textural features [40] are a total of 14 features calculated using a Grey Level Co-occurrence Matrix (GLCM). GLCM is used for texture analysis because it can estimate image quality related to second order statistics. The grey level co-occurrence matrix is a two dimensional matrix of joint probabilities between pairs of pixels separated by a distance *d* in a given direction *r*.

GLCM texture captures the relationship between two pixels at a time, known as the reference pixel and the neighbor pixel. GLCM shows the distance and angular spatial relationship over a specific size image sub-region. It considers how often the values of a pixel with a grey level (grey scale intensity or grey tone) are levelled horizontally, vertically and diagonally. GLCM is effective for the calculation of haralick features if the images are of same resolution and are grey-scaled [41].

In our study, we have used only the first 13 of haralick features because of the instability of ‘maximum instability coefficient’ (14th feature). The distance *d* is fixed at 1 and the direction *r* is varied as 0°, 45°, 90°, 135°. Mahotas [42], a computer vision python library, has been used to calculate the Haralick features in all the above mentioned directions. From the given dataset, each image is resized to 224 × 224 pixels and converted to a grey scaled image, to get the images into same resolution and and use grey levels, which is then passed through mahotas and a feature vector of 13 × 4 is extracted for each image.

### 2.4. Classification Using Random Forest

Random forest (RF) is an ensemble (i.e., a collection) of tens or hundreds of decision trees [43]. Ensemble models are often robust to variance and bias. The algorithm is efficient with respect to a large number of variables since it repeatedly subsets the variables available. Consequently, the deep spatial features generated by the auto-encoder and the spatial-temporal features produced by GLCM are concatenated and fed into the RF for classification.

### 2.5. Parameters of results assessment

For the assessment of the results obtained from the above process, certain quantities need to be evaluated which are widely used for comparison of the performance of binary classification models. In the given equations, follow the notation: TP be true positive, FP be false positive, FN be false negative, and TN be true negative.

1. Accuracy - It is used to convey the percentage of images correctly identified. It is defined as the ratio of correct predictions to the total number of predictions and is given as

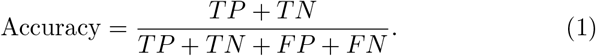
2. Precision - It is used to convey how many predicted as positive by the model were actually supposed to be predicted as positive. It is defined as the ratio of predicted true positives to the total number of predicted positives, and is given as

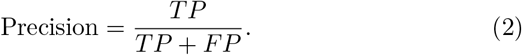
3. Recall - It is used to convey how well the model has classified the positive examples. It is defined as the ratio of predicted true positives to the actual positives, and is given as

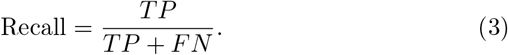
4. F1-score - It is the harmonic mean of precision and recall whose value is high if precision and recall are close and vice versa. It conveys how well the model is fitted, and is given as

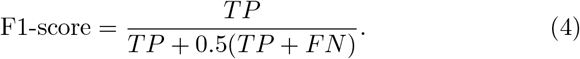
5. Specificity - It is used to convey how many predicted negative by the model were actually supposed to be predicted as negative. It is defined as the ratio of predicted true negatives to the total negatives predicted by the model, and is given as

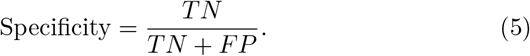
6. Area Under Curve - AUC conveys how well the model can distinguish between the positive and the negative class of the data.

## 3. Results and Discussion

Applying the proposed procedure on the input, two new datasets are generated, that is, one with features retrieved using autoencoders and other from Haralick features. From these datasets, a composite dataset is formed that contains the combination of these features as shown in Figure 2. A total of 564 relevant features are obtained which are used to train a random forest model with ntree = 100.

**Figure 2:**
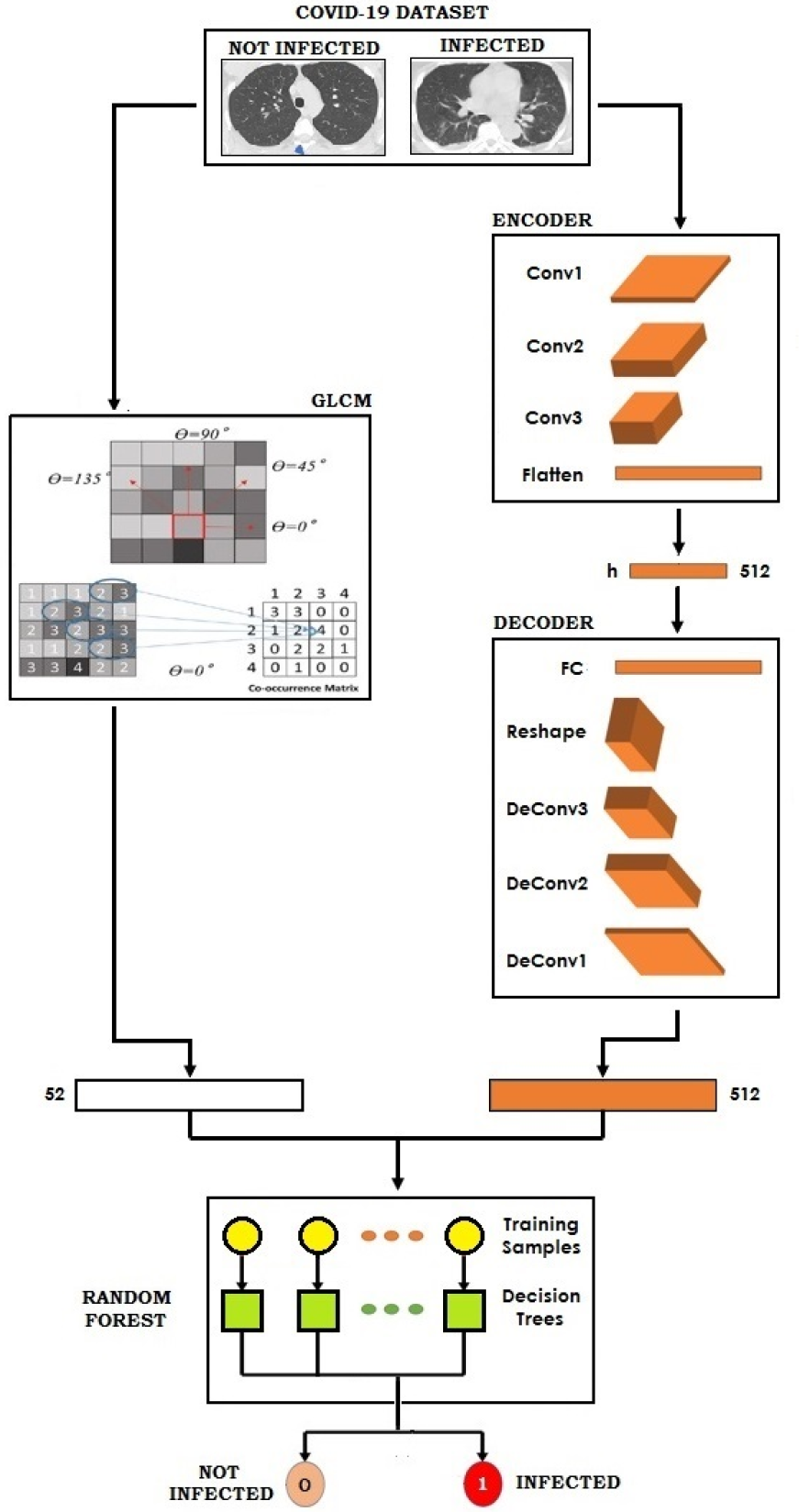
Diagrammatic representation of entire process

The input dataset was split into 80% training and 20% testing, that is, 1984 training and 497 testing images. Consequently, confusion matrices were generated for the classification models given in Table 2. This helps us to realise whether the model is confusing between two classes or not. Each row of the matrix represents the instances in a predicted class, while each column represents the instances in an actual class.

**Table 2:** Confusion matrices of dataset formed at each stage for 497 number of testing data points, where AP refers to Actual Positive, AN is Actual Negative, PP is Predicted Positive, and PN is Predicted Negative.

| GLCM |  |  | Autoencoder |  |  | Composite |  |  |
| --- | --- | --- | --- | --- | --- | --- | --- | --- |
|  | *AP* | *AN* |  | *AP* | *AN* |  | *AP* | *AN* |
| *PP* | 236 | 12 | *PP* | 234 | 14 | *PP* | 245 | 3 |
| *PN* | 11 | 238 | *PN* | 26 | 223 | *PN* | 8 | 241 |

Using the values obtained from the confusion matrices, the assessment of the model is done by calculating the performance metrics, summarized in Table 3.

**Table 3:** Performance metrics for each method

|  | GLCM | Convolutional encoder | **Composite** |
| --- | --- | --- | --- |
| Accuracy | 95.37% | 91.95% | **97.78%** |
| Precision | 95.2% | 94.09% | **98.77%** |
| Recall | 95.58% | 89.55% | **96.78%** |
| F1-score | 95.39% | 91.76% | **97.77%** |
| Specificity | 95.2% | 94.09% | **98.77%** |
| AUC | 95.37% | 91.95% | **97.78%** |

The column marked in bold is the proposed model which performed relatively better. Furthermore, the superior classification ability is also well demonstrated by Figure 3 and Figure 4 which are illustrations of prediction variability.

**Figure 3:**
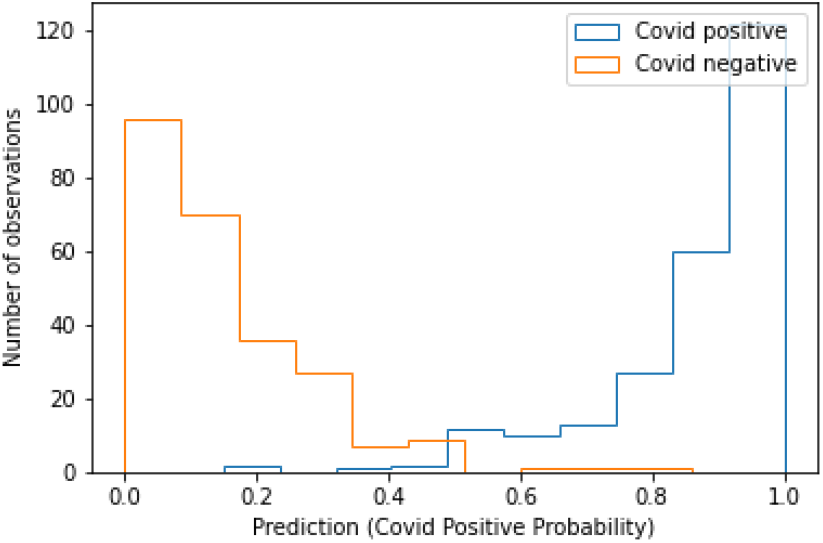
Plot of number of observations v/s prediction probability

**Figure 4:**
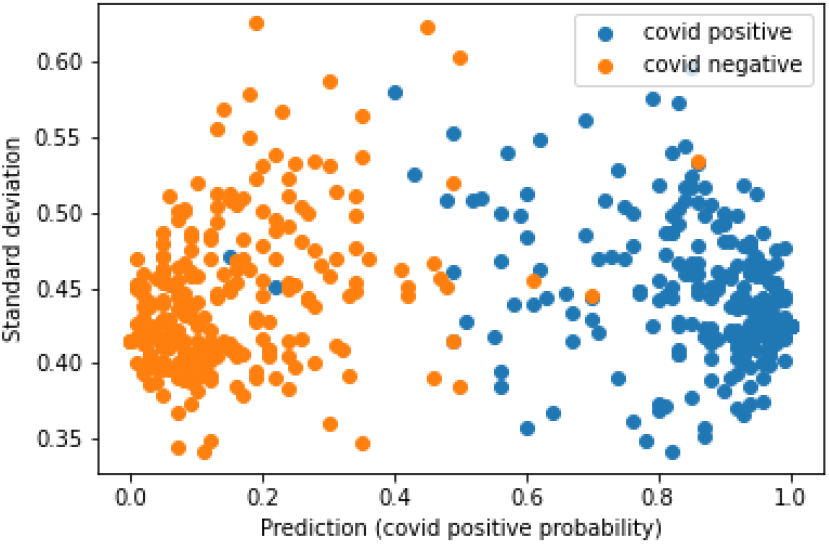
Scatter plot of standard deviation v/s prediction probability

These figures show the degree of influence the training set has for producing the observed random forest predictions and provides additional information about prediction accuracy.Forest-confidence-interval is a Python module for calculating variance and adding confidence intervals to scikit-learn random forest classification objects The package’s *random_forest_error* function uses the random forest object (including training and test data) to create variance estimates that can be plotted (such as confidence intervals or standard deviations).

The classification accuracy, F1-score, precision, recall, specificity and AUC for the developed framework is 97.78%, 97.77%, 98.77%, 96.78%, 98.77% and 97.78%, respectively. The time taken by the model to predict 50 training examples is 13.4 seconds, that is, 0.268 seconds per testing image. Thus, the results demonstrate that the fusion of texture features with deep feature can provide a representative description for COVID-19 detection in a fast and efficient manner. We also performed the whole analysis with support vector machine as a classifier but its performance was poor as compared to random forest based model. We can therefore conclude that the proposed model is indeed a better model with discriminatory features for the classification of COVID-19 from CT images.

## 4. Conclusion

In this paper, we proposed a new method of extracting features, extracted from the CNNs autoencoder with powerful handcrafted GLCM feature combine with traditional machine learning algorithm, to efficiently handle data complexity in order to accurately predict COVID-19 from chest CT images. Thus we are able to show that the two paradigms of the extraction feature are able to extract information which the other paradigm neglects. This finding may have a significant impact on the treatment of COVID-19 patients, because it is simple and reliable, thus improving early detection. The success of the proposed model supports the representative power of CNNs and traditional machine learning approach. However, with regards to the use of either traditional machine learning models or CNN classifiers, the new approach has considerable advantages in terms of accuracy.

## Data Availability

www.kaggle.com/plameneduardo/sarscov2-ctscan-dataset

## Data availability

The data that supports the findings of this study is available at www.kaggle.com/plameneduardo/sarscov2-ctscan-dataset.

## Author Contribution Statement

C.G. majorly conducted experimentation and evaluation/compilation of results. The writing and organization of the manuscript was primarily handled by A.K., conception and design, and critical review by S.K.D., and later the paper was thoroughly read by V.S. who modified the presentation of the paper’s contents and the results section. The methodology section was jointly contributed by all.

